# An ensemble deep learning model with empirical wavelet transform feature for oral cancer histopathological image classification

**DOI:** 10.1101/2022.11.13.22282266

**Authors:** Bhaswati Singha Deo, Mayukha Pal, Prasanta K. Panigrahi, Asima Pradhan

## Abstract

Oral squamous cell carcinoma (OSCC) has become quite prevalent across many countries and poor prognosis is one of the major reasons for the ensuing high mortality rate. It mainly occurs in sites such as tongue, tonsil, oropharynx, gum, floor and other parts of the mouth. For early detection, the widely used approach is biopsy, in which a small portion of the tissue is taken from the mouth and examined under a disinfected and secure microscope. However, these observations do not effortlessly distinguish between normal and cancerous cells. Diagnosis of OSCC is generally done by pathologists who mostly rely on their years of empirical experience from tissue biopsy sections. The possibilities of human errors increase while detecting the cells using microscopy biopsy images physically. With the growth of artificial intelligence, deep learning models have gained immense importance in recent years and have become one of the core technologies in numerous fields including the prediction of lung cancer, breast cancer, oral cancer, and various medical diagnosis. It not only enhances accuracy, but also fastens the image classification process, as a result, lowering human errors and workload. Here, we have made use of a customized deep-learning model for aiding pathologists in better OSCC detection from histopathological images. We accumulated and analyzed a complete set of 696 histopathological oral images, amongst them 80% have been taken in the training set, 10% of the images are included in the validation set, and the rest 10% for testing purposes. In this study, 2D empirical wavelet transform is used to extract features from the images; later an ensemble of two pre-trained models, namely Resnet50 and Densenet201 are used for the classification of images into normal and OSCC classes. The efficacy of the model is assessed and compared in terms of accuracy, sensitivity, specificity, and ROC AUC scores. The simulation results show that the proposed model has achieved an accuracy of 92.00%. Thus, this method may be utilized for assisting in the binary classification of oral histopathological images.

## 1. Introduction

Oral cancer is one of the most prevalent fatal diseases and has long been a major public health problem in countries across the globe. More than 90% of oral cancers are oral squamous cell carcinomas (OSCC), a diverse group of cancers that emerge from the mucosal lining of the mouth cavity (1). According to the World cancer fund research international (2), cancers of the lip and oral cavity are the most prevalent, with more than 377,700 cases reported worldwide in 2020, being the 11th and 18th most prevalent types of cancer, respectively, in men and women (3). The most common indication of cancer is an ulcer or sore that does not cure and may cause pain or bleeding. Unhealing white or red sores on the lips, tongue, gums, or cheeks, a mass or lump in the mouth, swallowing or chewing issues, jaw swelling, difficulty in speaking and chronic throat pain are common symptoms of oral cancer. Reasonably carcinogenic oral lesions are oral mucosal characteristics that have a higher chance of developing in to cancer than healthy mucosa. Risk factors include alcoholic beverages, tobacco use, poor dental hygiene, exposure to the human papillomavirus (HPV), as well as hereditary factors, mode of living, ethnicity, and location. Avoiding risk factors and boosting protective factors are both important aspects of cancer prevention. It is of paramount importance to detect oral cancer early, most people are diagnosed when it is well advanced, which leaves them with a poor prognosis. The clinical strategy for treating cancer and the way it affects a person’s quality of life and perception of competence for day-to-day functioning are reflected by therapy.

Early OSCC identification is crucial for a successful course of treatment, improved survival rate, and low morbidity and mortality rates (4). The OSCC has a poor prognosis with an average cure rate of 50% (5; 6). The standard technique for diagnosing OSCC is histological examination of tissue samples under a microscope (7; 8). Typically, histopathological images are viewed at a smaller magnification level allowing extensive considerable inspection at the tissue level. The distribution of cells, cell size and shape, and size and form of the cell nucleus are the main criteria used to identify cancer cells. Before performing surgery, histopathological analysis is crucial to determine the tumor size as precisely as feasible. This diagnostic pathology approach relies on the interpretation of histopathologists, which is often laborious and prone to mistakes, limiting its clinical applicability (9). As a result, it is significant to equip pathologists with efficient diagnostic tools to assist in the evaluation and diagnosis of OSCC.

Recent years have seen an increase in research on using artificial intelligence (AI) to enhance medical diagnosis. Scientists from all over the world are fascinated by artificial intelligence, a technological advancement that mimics human cognitive abilities (10; 11). Artificial intelligence lessens the pressure on doctors, simplifies their workload, and relieves their exhaustion (12). Due to the rise in diagnostic imaging usage, researchers have been able to investigate AI applications in medical image processing (13). Particularly in the diagnosis of pathological images, deep learning (DL) has demonstrated excellent effectiveness in resolving a number of medical image processing difficulties (14; 15; 16). There have been a number of studies done on the analysis of medical images through smartphones that use artificial intelligence-based algorithms. Recently, AI has been used in dentistry, and the results are astounding. Oral cancer patients can now more easily be identified, treated, and monitored due to artificial intelligence technology (17). Artificial intelligence-identified biomarkers from numerous studies have revealed prognostic factors for oral cancer.

Deep learning approaches have generated a lot of interest in recent years due to their applicability and purported advantages in the field of cancer prediction. This has helped doctors in encouraging and enhancing better patient health management. Deep neural networks have overtaken neural networks as a result of technical improvements. For a diverse range of cancer types, namely breast cancer, prostate cancer, and lung cancer, large-scale computer-aided diagnostic (CAD) systems based on DL have been proposed and developed (18; 19; 20). CAD systems are becoming more effective and sophisticated due to breakthroughs in machine learning (21). These tools are necessary for quicker, more accurate analysis of the histopathological images and the detection of irregularities. In order to accurately diagnose cancer cells, strong pre-processing techniques and learning algorithms are needed since histopathological images provide more information about the structural attribute of the underlying tissues. A detailed study was conducted to examine the goal of examining the possible applications of deep learning in oral cancer in the context of the prevalent characteristics of this disease, including the growth in occurrence, the need to improve diagnostic tools, and the amount of literature on its usage.

## 2. Related works

This section reviews several cslassification methods for oral cancer in two categories. In the first and second categories, machine learning-based approaches and deep learningbased approaches are discussed respectively.

### 2.1. Machine Learning based methods

The majority of research focuses on using machine learning techniques to identify oral submucous fibrosis (OSF) (22; 23; 24). As OSF grows, its chronic nature could result in oral cancer. A technique for “Textural characterization of histopathology images for oral sub-mucous fibrosis diagnosis” was put forth by Muthu et al.(25). They combined various wavelet family characteristics with an SVM classifier to improve accuracy. Using run-length and texture features, T.Belvin et al. (26) employed backpropagation-based ANN to classify oral lesions into multiple categories. The suggested model uses a mix of GLCM and GLRL characteristics in order to achieve a more illustrative and patientspecific approach. T. Y. Rahman et al (27; 28) conducted studies that are centered on the binary classification of OSCC. According to these investigations, texture, shape, and color feature allowed for 100% classification accuracy using Support Vector Machine (SVM) and Linear Discriminant Classifier (LDA). Numerous patterns and behaviors can be seen in oral cancer (29). In recent years, researchers have utilized various machine learning techniques to detect and overcome cancer (30). Much better than prior predictions, machine learning can now predict oral cancer methods (31). To combat cancer, researchers can employ a variety of histopathological machine learning techniques (32). The works mentioned above represent contemporary efforts to predict and diagnose oral cancer employing machine learning methodologies.

### 2.2. Deep Learning based methods

Deep learning techniques, particularly convolutional neural networks (CNNs), have become the state-of-theart method for many tasks involving image analysis (33). By performing feature extraction and classification tasks simultaneously, CNN with deep learning performs well for image problems. Due to the advent of powerful GPUs, deep learning methods can classify histopathological images with greater accuracy without manually representing the features of the input data (34). In order to classify four different types of OSCC, Navarun et al. (35) used four pre-trained models through transfer learning and compared them with a suggested CNN model. For the categorization of oral cancer tissue into seven classifications, Jonathan et al. used Random Learning (RL) and Active Learning (AL) through CNN (lymphocytes, stroma, adipose, tumor, keratin pearls, mucosa, and blood). It was discovered that the AL’s accuracy was 3.26 percent more than the RL’s (36).

The purpose of study (37) was to investigate state-of-theart automated methods for detecting OSCC on clear images utilizing extensive training and CNN techniques. This CNN focuses on finding quotes, images, training, data, and grading. Even though several machine learning-based studies on oral cancer have been proposed, relatively little work has been done utilizing deep learning to analyze histopathological images. The majority of earlier studies used deep learning and machine learning models to detect oral cancer utilizing OSCC biopsies or other datasets, however, they were unable to achieve the highest accuracy due to its noted limitations. As previous studies have demonstrated, predicting oral cancer is a crucial goal to save many lives. In this work, an efficient feature extraction strategy is proposed that combines an empowered transfer learning model with empirical wavelet transform to extract features and train on OSCC histopathological images to detect oral cancer.

### 2.3. Research gaps and motivation

1. Convolutional neural networks have made numerous advances in the field of image classification (38; 39; 40). However, they are challenging to train for two reasons. First, the initial layers train relatively slow because of the gradient’s exponential decline. Second, it takes longer to train CNN models since they include more parameters that increase the network’s complexity.
2. Although batch normalization and activation functions have made significant advancements in reducing the impact of exploding/vanishing gradients, optimizing a neural network with intricate architecture is still difficult (41). We have proposed an ensemble method to classify oral cancer histopathological images into two groups in order to address this concern.
3. Machine learning-based approaches, in contrast to deep learning approaches, demand domain knowledge to recognize handcrafted features from histopathological images for efficient categorization.
4. In majority of the published research works, class imbalance has been noted in the datasets utilized for training the deep learning models. It will be difficult to generalize the feature patterns for all classes given this imbalance. As a result, when training deep learning models, appropriate and efficient augmentation approaches must be applied.

### 2.4. Research contributions

Listed below are the primary contributions of this work:

1. We proposed a method in which feature extraction is carried out employing 2D empirical wavelet transform and further histopathological images are classified into normal and OSCC classes using a weighted ensemble of Resnet50 and Densenet201 architectures. This method is effective by improving accuracy and maintaining information across layers.
2. The proposed ensemble method architecture offers improved performance with a limited dataset by converging more quickly and requiring less computing complexity.
3. It incorporates considerable data augmentation to increase the amount of training data in order to address the issue of class imbalance. This made the proposed network more reliable and prevented overfitting.
4. For categorization, the majority of research studies used a single-path deep learning architecture. It was noted that these networks had slightly higher false positive and false negative rates. By using cutting-edge techniques like feature fusion from different tracks, ensembling, etc., this can be resolved.

The remaining part of this work is presented as follows: section “Proposed Method” discusses details of the 2D empirical wavelet transform, Resnet50 and Densenet201 architecture, transfer learning, and ensemble learning, section “Data and materials” includes a description of the dataset, preprocessing, training criteria, data augmentation techniques, and implementation, section “Results and Discussion” discusses the evaluation metrics, hyperparameters used, and results of the proposed method. Finally, the section “Conclusion” presents the conclusion and upcoming works.

## 3. Proposed Method

In this work, we have proposed a two-dimensional empirical wavelet transform (2D-EWT) based feature extraction approach on the histopathological images to generate subband images. We have considered the first subband image in our training process. The augmented subband images are then taken as input to Resnet50 and Densenet201 pre-trained models for training. Finally, to enhance the performance of the model, the weighted ensembling technique is used followed by accuracy evaluation on the test dataset. Figure 1 shows the schematic diagram of the feature extraction method employing EWT. Figure 2 shows the first three subbands of a sample oral cancer histopathological image. Figure 3 shows the oral cancer histopathological image classification framework. Figure 4 layers of the convolutional neural network used in the Resnet50 and Densenet201 model.

**Figure 1:**
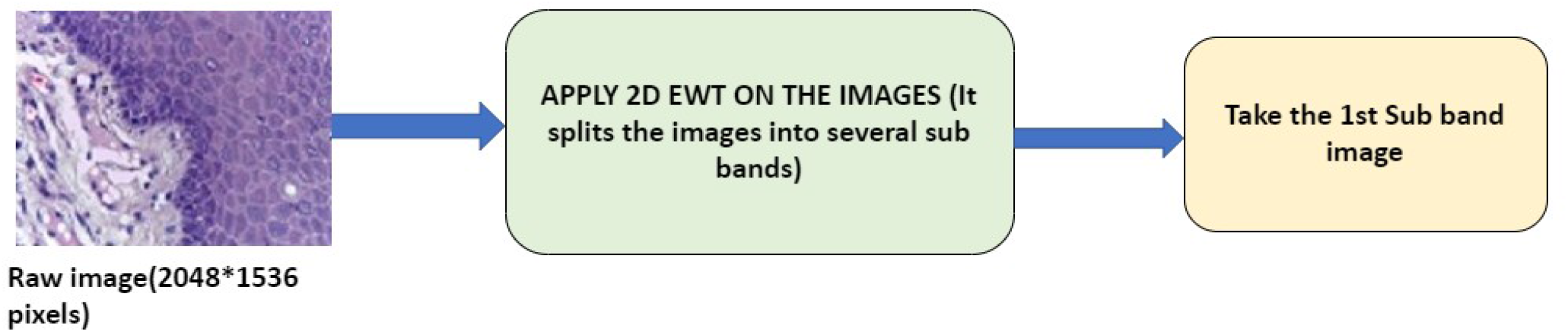
Methodology for feature extraction using Empirical wavelet transform for oral cancer histopathology image classification

**Figure 2:**
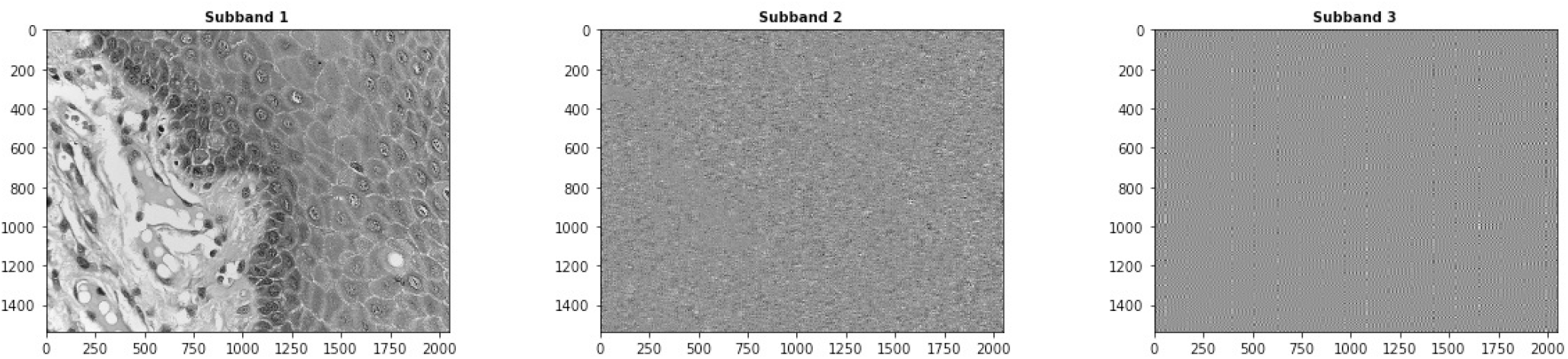
First three subbands of a sample oral cancer histopathology image

**Figure 3:**
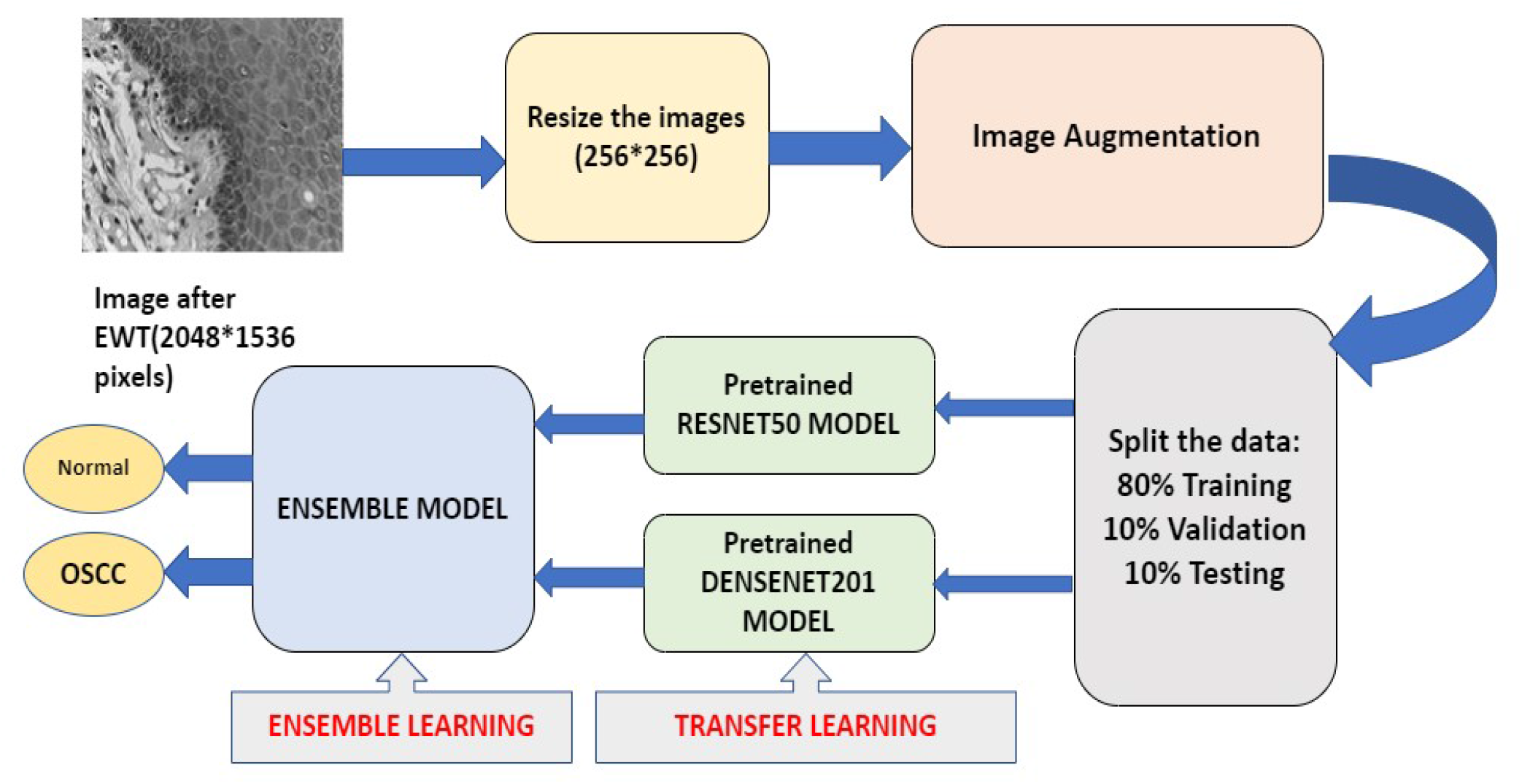
Proposed methodology for the classification of oral cancer histopathological images using ensemble deep learning with empirical wavelet transform

**Figure 4:**
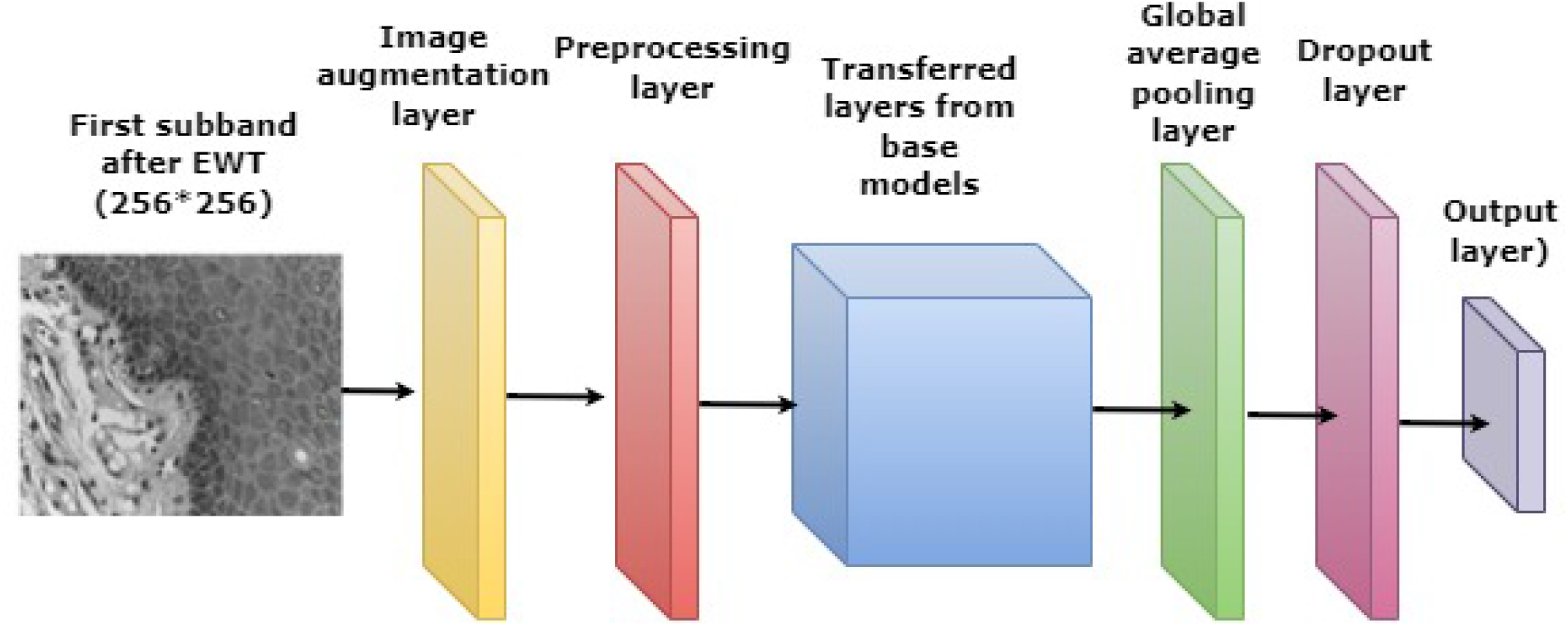
Convolutional neural network of Resnet50 and Densenet201 model

### 3.1. 2D Empirical wavelet transform

For the purpose of signal processing, Gilles (42) presented the empirical wavelet transform (EWT). Gilles expanded it in 2014 to allow for the use of EWT with image domain (43). It is evident from the research that EWT has a variety of uses, including estimating power quality indices of power signals (44; 45), estimating pitch (46), predicting wind speed (47), and diagnosing bearing faults (48). A cross-term decline in the Wigner-Ville distribution and automatic coronary artery disease diagnosis have both been studied using the EWT approach (49; 50). Different signal decomposition techniques have been researched to break down physiological signals including electromyograms, electrocardiograms, and electroencephalograms (EMG). Eigenvalue decomposition (EVD), Multivariate EMD (MEMD), Empirical mode decomposition (EMD), EWT, and 2D EWT are few of them (51; 42; 43).

In (42), the researcher proposed formulating an empirical wavelet transform (EWT). The concept includes creating a collection of M wavelet filters with (M-1) bandpass filters and one lowpass filter related to the details and approximation components, respectively. These filters are centered on “well-chosen” Fourier supports (meaning relevant modes are selected in the signal spectrum). If *f* (*t*) is a 1D signal and its fourier transform is denoted by |*F*_1,*t*_ (*f*) | (*ω*), then we first use |*F*_1,*t*_ (*f*) | (*ω*) to identify the boundaries of each Fourier support (43). We are given a set of boundaries via this operation Ω = {*ω*_*n*_}_*n*=0,…*N*_ (we limit our study to the range [0,*π*] and use the assumption that *ω*_0_ = 0 and *ω*_*N*_ = *π*). As in (43), we can create a wavelet tight frame, 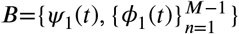 inspired by Meyer’s and Littlewood-Paley wavelets. The transition phase is centered around *ω*_*n*_ which has width of 2*γω*_*n*_ where 0<*γ*<1. Empirical scaling function is given as:

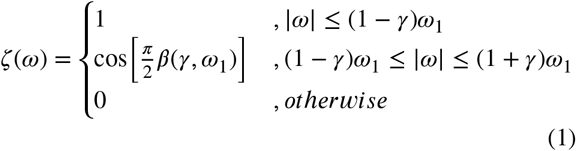

and empirical wavelet function is given by:

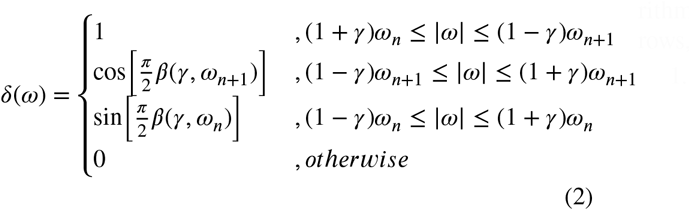

where *β*(*γ, ω*_1_),*β*(*γ, ω*_*n*_) and *β*(*γ, ω*_*n*+1_) are represented by:

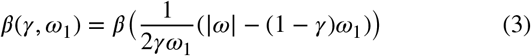

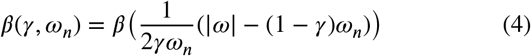

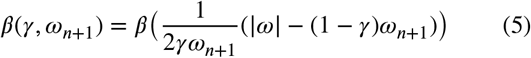

Also *β*(*z*) fulfills the conditions:

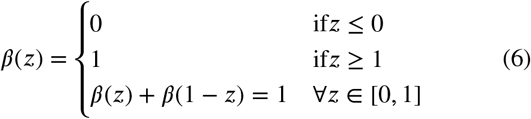

The tight frame property of B is given a necessary condition in (42), which enables us to arbitrarily choose this value. By taking into account a tensor product technique similar to the conventional 2D discrete wavelet transform, the researcher proposed extending the 1D EWT to images (43). It is intended to employ the 1D EWT on rows and columns individually. It is simple to understand how distinct sets of filters defined on certain different Fourier supports can be obtained if we consider each row (or column) separately. Algorithm: 2D Tensor Empirical Wavelet Transform algorithm. Let the image be *f* (*x*), *N*_*row*_ denotes the number of rows, and *N*_*column*_ denotes the number of columns (42).

1. Perform 1D FFT of each rows *i* of *f*; *F* (*i, ω*); and compute the average row spectrum magnitude.

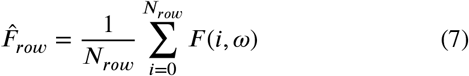
2. Perform 1D FFT of each columns *j* of *f*; *F* (*ω, j*); and compute the average column spectrum magnitude.

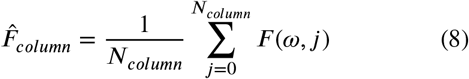
3. Detect the boundaries on 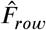 and construct the corresponding filter bank 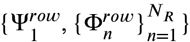.
4. Detect the boundaries on 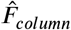 and construct the corresponding filter bank 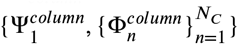.
5. With the filter bank 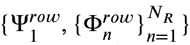, filter along the rows which gives *N* _R_ output images.
6. With the filter bank 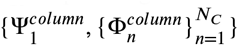, filter along the columns which finally gives (*N*_*R*_ +1) (*N*_*C*_ + 1) subband images.

where, *N*_*R*_ and *N*_*C*_ are average row and column subbands respectively.

### 3.2. Resnet50 Architecture

A classic deep convolutional neural network called ResNet integrates classification, auto-encoding, and images. It utilised feature transmission to minimize vanishing gradient problem and went on to win the 2015 ImageNet Large Scale Visual Recognition Challenge (ILSVRC) by enabling the training of considerably deeper networks than those previously employed (52). It is well known that stacking layers on top of each other does not work in efficiently increasing the network depth. Due to the vanishing gradient problem, the gradient in deep networks may become very small when gradients are repeatedly multiplied as they are back-propagated to prior layers, As a consequence, when the network gets deeper, its performance becomes saturable or even begins to decline quickly.

Skip connection was first introduced by ResNet. The skip connection is depicted in figure 5. Bypassing some intermediate levels, the skip connection links activations from one layer to the next. Consequently, a block is left over. To build resnets, these leftover blocks are piled. Instead of having layers learn underlying mapping, the approach used by this network is to allow the network fit the residual mapping. This type of skip link has the advantage of allowing regularisation to bypass any layer that impairs architecture performance. As a result, disappearing or increasing gradients are not a problem while training an exceptionally deep neural network.

**Figure 5:**
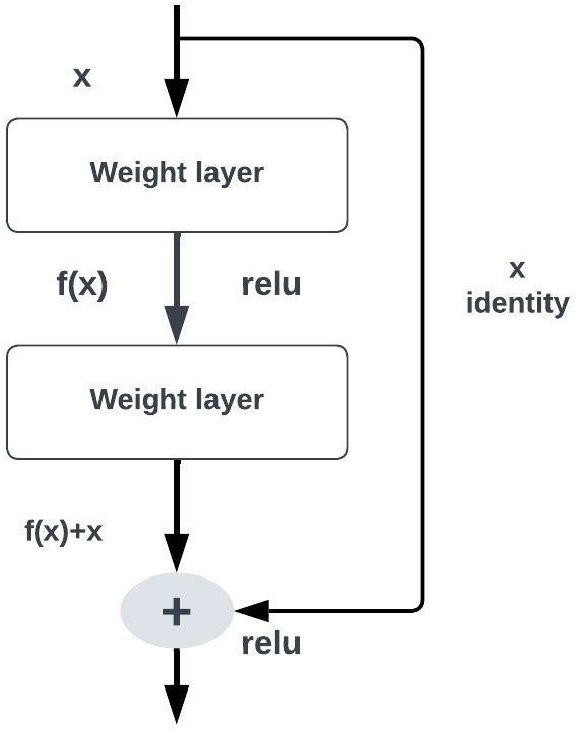
Structure of Resnet block showing the skip connection

### 3.3. DenseNet201 Architecture

Each layer in DenseNets by Huang et al. (53) is made up of the feature maps of the present layer and all former layers. As a result, these networks are efficient in terms of computation and memory usage, have rich feature representation for the input images, and are compact (i.e., have fewer channels). If there are shorter connections between the layers closest to the input and the layers closest to the output, CNN can be trained more quickly, be significantly deeper, and be more accurate. The dense convolutional network (DenseNet) has a feed-forward connection between each layer and every other layer. The feature maps of all the preceding layers are used as input for each layer, and the feature map of the current layer is utilized as input for all subsequent layers as shown in figure 6. The vanishing gradient problem is avoided and its effects are lessened due to DenseNet, which also strengthens feature propagation, encourages feature reuse, and drastically reduces the number of parameters.

**Figure 6:**
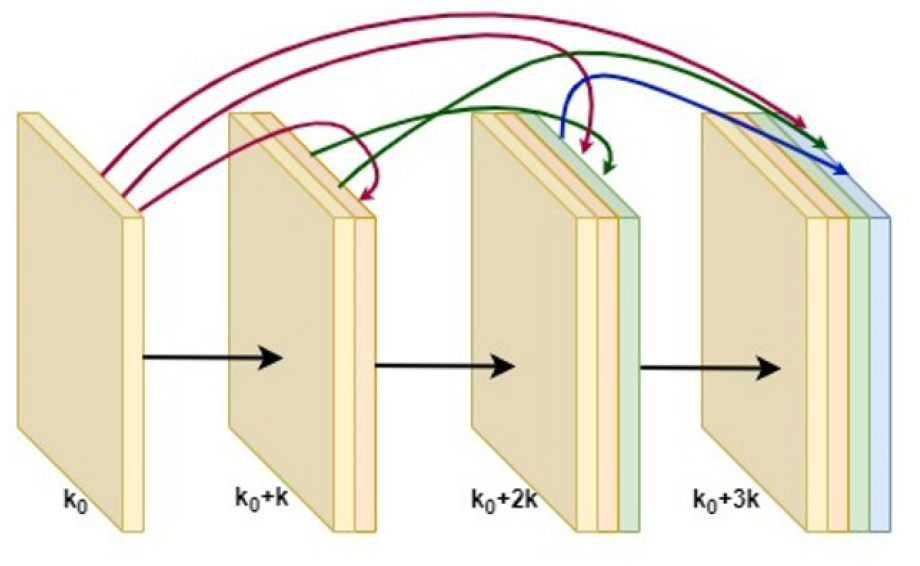
Structure of Dense block

### 3.4. Transfer Learning

Medical imaging, a powerful tool for diagnostics, plays a significant role in the medical field. Computer-aided diagnosis has grown in popularity and potential with the advancement of computer technologies such as machine learning. Keep in mind that medical images are produced with specialized equipment, and their labeling frequently relies on skilled physicians. Therefore, gathering enough training data is frequently expensive and challenging. Medical imaging analysis can make use of transfer learning technology. Transfer learning is the process of using features discovered while solving one problem to solve another that is somewhat related. Pretraining a neural network on the source domain such as ImageNet, a collection of more than 14 million annotated images with more than 20,000 categories (54), and then refining it using examples from the target domain is a typical transfer learning technique. Similar to this, Shin et al. improved the pre-trained deep neural network to address the issues with computer-aided detection. In order to assess knee osteoarthritis, Byra et al. (56) used the transfer learning technique. Transfer learning has some more uses in the field of medicine besides imaging analysis. For instance, Tang et al. study (57) integrates domain adaptation and active learning technologies for the classification of varied medical data. In order to automatically encode ICD-9 codes that characterize a patient’s diagnosis, Zeng et al. (58) applied transfer learning.

Pre-trained Resnet50 and DenseNet201 models are used in the proposed method, and it is trained with new data using transfer learning as shown in Figure 7. The workflow followed in the process of transfer learning is:

**Figure 7:**
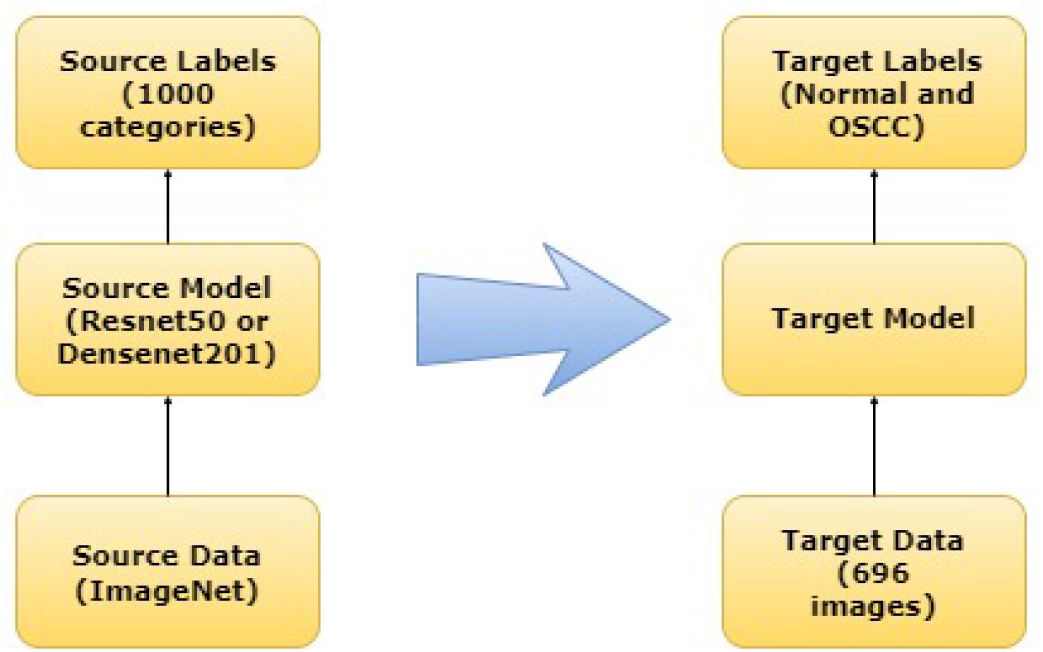
Transfer learning framework

1. Use layers from a model that has already been trained.
2. Freeze them to prevent losing any of the data they contain during upcoming training rounds.
3. On top of the frozen layers, add some fresh, trainable layers. On a new dataset, they will discover how to forecast using the previous features.
4. Utilizing the dataset, train the new layers.

### 3.5. Ensemble Learning

There is a lot of evidence that an ensemble of models, which is a combination of separate models that can be merged to reduce variance, bias, or both, can perform better than a single model, such as a single decision tree, in terms of making numerous predictions (59). A single model is unlikely to accurately capture the underlying features of the data in order to generate the most accurate predictions. This is where combining several models can significantly increase prediction accuracy. Multiple deep learning models could be combined to extract more features from the underlying structure of the data (60; 61). The Kaggle competitions, the Data Mining World Cup, and Netflix Prize are just a few examples of real-world uses for ensemble modeling (62; 63; 64).

Even though ensembling models are highly popular in data analytics, not all ensembles are created in the same fashion. Different ensembling techniques include boosting, bagging and stacking/blending (65; 66; 67). Boosting creates an ensemble by integrating weak learners in the possibility that later models will correct mistakes made by previous models (68). Using replacement sampling (bootstrap), bagging generates an ensemble by voting or averaging across class labels on the training dataset (65). Stacking uses a different learning method to determine the response values in comparison to the results of the basic learners on the training dataset (69). Each approach has advantages and disadvantages. Bagging does not perform well with relatively basic models and tends to lower variance more in comparison to bias. Boosting seeks to lessen bias and variation by merging weak learners in a sequential manner, but it is vulnerable to noisy data, outliers, and over-fitting. While stacking aims to reduce variance and bias by likely fitting multiple meta-models to the predictions provided by base learners, hence correcting the mistakes made by base learners. We emphasize stacking with a weighted average in this work, which involves integrating base learners with a weighted average as shown in figure 8.

**Figure 8:**
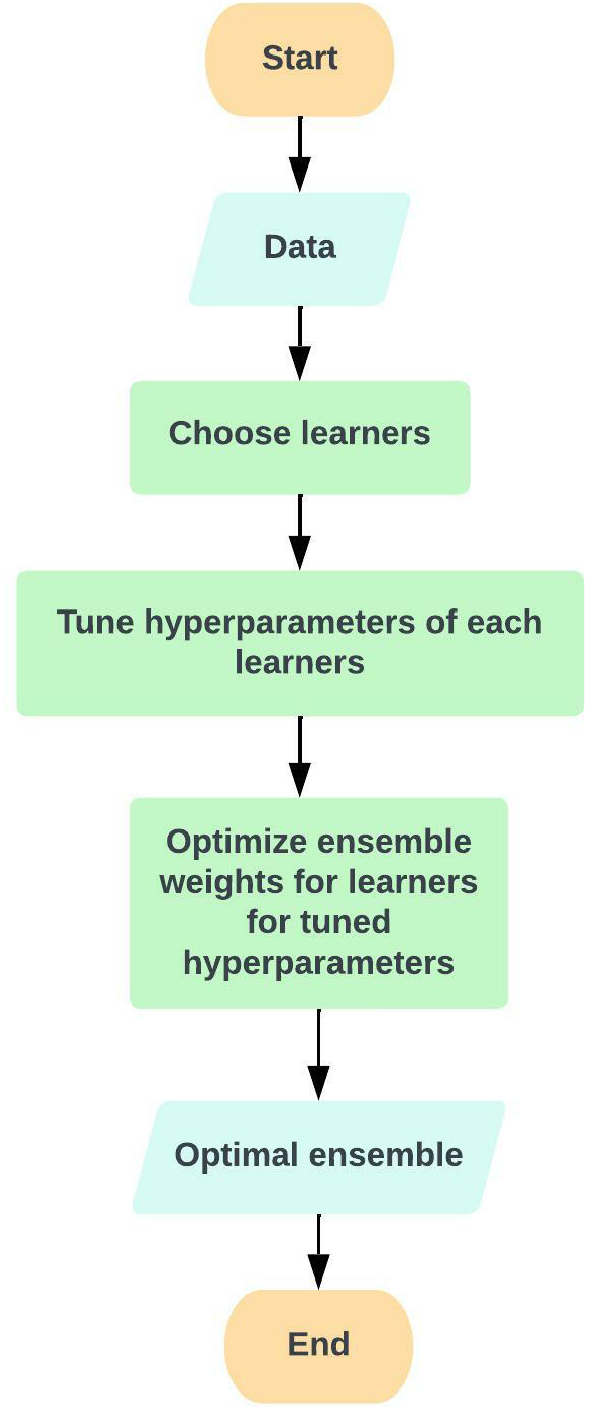
Weighted ensemble flowchart

## 4. Data and materials

This section introduces the dataset description, preprocessing, training criteria, data augmentation techniques, and implementation details.

### 4.1. Dataset Description

Hematoxylin and eosin (H&E) stained microscopic images with 400x magnification level are retrieved from the first set in (70), which is publicly accessible. There are two categories of patients in this oral cancer dataset: those who have oral cancer and healthy patients. This dataset is used by the proposed model to evaluate and predict oral cancer. Figure 9 and 10 display samples from both categories of the dataset, while table 2 displays the complete dataset classes.

**Table 1.**
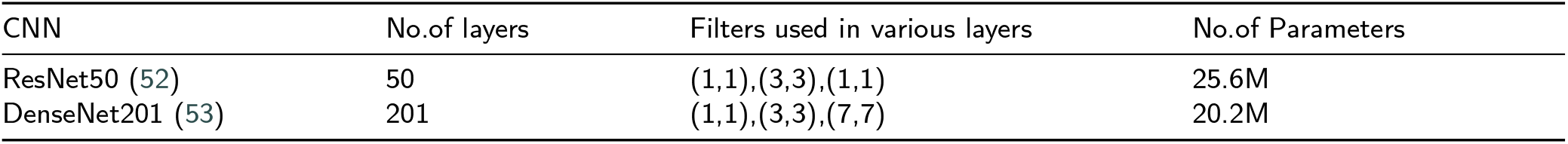
Number of layers and parameters in each network

**Table 2.** OSCC biopsy data instances

| Classes | Images |
| --- | --- |
| Normal | 201 |
| OSCC | 495 |

**Figure 9:**
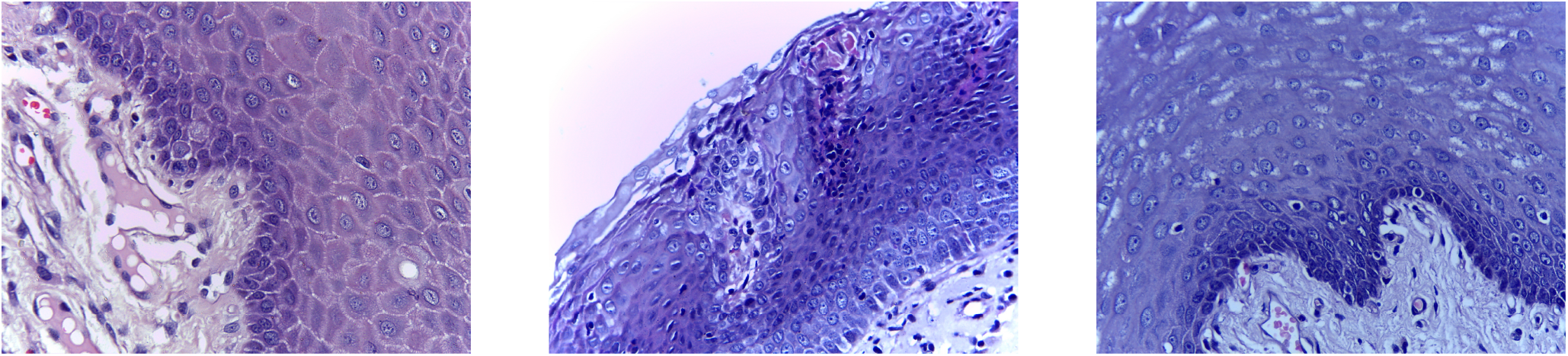
Sample Images from the Normal subjects

**Figure 10:**
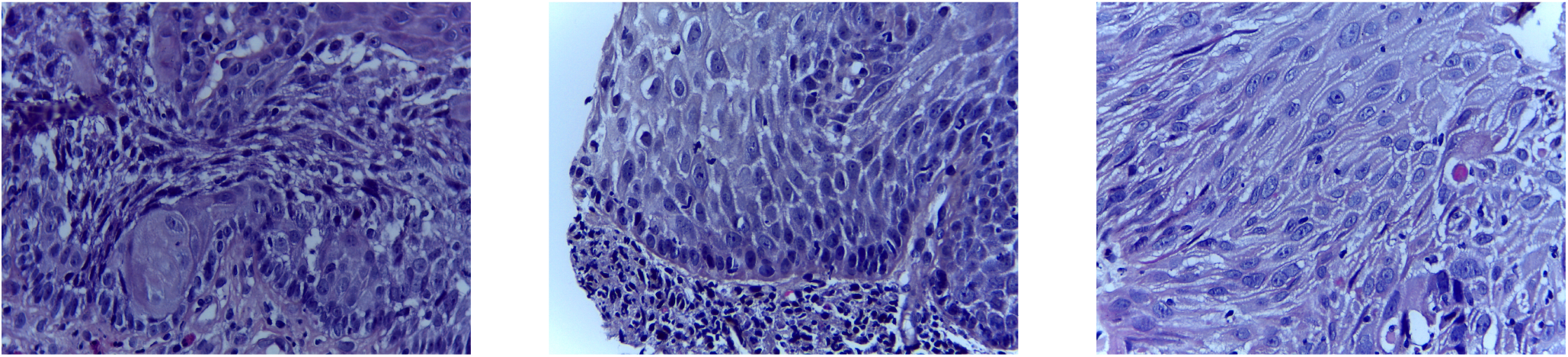
Sample Images from the OSCC patients

### 4.2. Preprocessing

The dataset includes images with 2048×1536 pixels. The Image Data Generator library from Keras is used to reduce the size of the images to 256 × 256 pixels prior to training. This will have a major impact on learning generalised feature patterns and network convergence. Each input channel is normalized in relation to the ImageNet dataset and the pixel values of the input image are scaled between 0 and 1.

### 4.3. Training Criteria

For the Resnet50 and Densenet201 models, the percentage of normal and OSCC images was kept the same, 80% of the images were chosen for training, 10% of the images for validation, and 10% of the images for testing purposes. In this form, the model was trained on 576 images, validated on 69 images, and tested on 71 images. Table 3 displays this data regarding the models’ training, validation, and testing.

**Table 3.** Training, validation and test images criteria

|  | Images | Percentage |
| --- | --- | --- |
| Training | 596 | 80% |
| Validation | 69 | 10% |
| Test | 71 | 10% |

### 4.4. Data Augmentation

Image data augmentation is a method for increasing the dataset that involves creating altered images during the training phase. The class imbalance can lead to overfitting and poor convergence during training because the dataset has more samples of OSCC (71.12%) than normal images (28.88%). A set of augmented images must be generated in order to solve this issue. We employ the ImageDataGenerator function provided by the Keras deep learning toolbox to generate sets of tensor image data with data augmentation being done in real-time. Thus, we ensure that every time our model is trained, it sees new iterations of our image. The ImageDataGenerator receives a batch of input images and then randomly rotates, flips, standardizes, brightens, shifts, and alters each image in the batch. The images are flipped horizontally by setting “Random Flip = Horizontal”. One of the often-used strategies for augmentation is image rotation, and the ImageDataGenerator class randomly rotates images to a certain degree. Here setting “Random rotation = 0.2” specifies the image is being rotated in the range [-20% × 360°, +20% × 360°]. Additionally, we select “Random height = 0.2,” which specifies the maximum percentage of total height by which the image may be arbitrarily shifted upward or downward. Images are zoomed in between 20% to 30% by setting “Random zoom=0.2 to 0.3”. The calling function is then given the batch that has undergone a random transformation. Table 4 displays each of these variables along with their values.

**Table 4.**
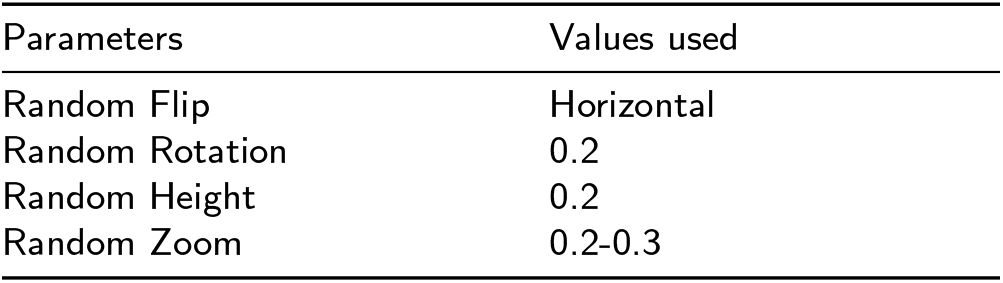
Parameters of Image Augmentation

### 4.5. Implementation

All experiments associated with this article are implemented on a normal PC with Intel(R) Iris(R) Xe graphics by using TensorFlow 2.7.0, Python 3.7.6, and Keras 2.7.0. Additionally, this computer contains a 2.40 GHz Intel(R) Core(TM) i5-1135G7 processor and 16.0 GB of RAM.

## 5. Results and Discussion

In this section, we discussed how to interpret the evaluation metrics for our proposed model, and later, we described how to tune the hyperparameters and finally discussed the results of Resnet50, Densenet201, and the proposed ensemble method by using EWT and without using EWT.

### 5.1. Evaluation Metrics

The efficacy of our proposed methodology highly depends on the information included in the confusion matrix, also known as the error matrix or contingency table. This confusion matrix contains four keywords: True Positive (TP), false positive (FP), false negative (FN), and true negative (TN). In our work, the TP indicates images that were accurately classified as OSCC, whereas the FP indicates images that were misclassified as normal. While the TN indicates accurately classified normal images and the FN indicates OSCC class images that were wrongly classified as normal. Precision, sensitivity, classification accuracy, and F1-score were used as performance measures based on the confusion matrix to analyze the classification efficacy of our proposed approach on the test images. These metrics were implemented using the Python scikit-learn module. These evaluation metrics can be computed using the formulas below:

1. Precision: The precision is determined as the ratio of positively classified positive samples to all positively classed samples (either correctly or incorrectly). The precision measures how accurately a sample is classified as positive by the model.

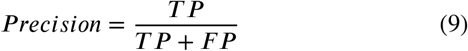
2. Sensitivity: Sensitivity, also called ‘recall’ determines the degree of completeness of a model. It displays the percentage of OSCC images that were correctly identified out of all OSCC images.

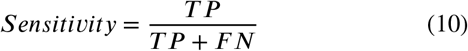
3. Accuracy: The fraction of correctly identified images to all test images, which assesses prediction performance.

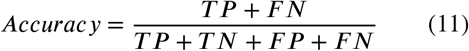
4. F1-Score: The F1 score is the harmonic mean of the model’s recall and precision. It is a technique for enhancing the model’s recall or precision.

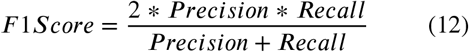

### 5.2. Hyperparameter tuning

Neural networks possess the potent ability to autonomously learn the relationship between their inputs and outputs (71). However, some of these correlations could be the result of sampling error, in which case they might be more prevalent throughout training but not in the actual test dataset. Such a challenge could lead to overfitting problems, which might impair the prediction performance of a deep learning model (71). To determine how well our proposed methodology performs generally, we strictly adhered to the hyperparameter tuning method. The following process was used to choose the best hyperparameters: First, for our binary classification problem, we chose binary cross-entropy as a loss function. Then, during the training procedure, the Adam (adaptive moment estimation) method (72) was utilized to carry out the optimization over 200 epochs. We explored three different learning rates (0.001, 0.0001, and 0.00001) and three different batch sizes (16, 32, and 64), taking into consideration the estimations used in other previously published papers. We found that a batch size of 32 and a learning rate of 0.001 worked well together to achieve our main model training goal of minimizing the generalization gap between training loss and validation loss. In addition, we used a 0.2 dropout to prevent the model from becoming overfitted during training (73). Then, based on which model had the lower validation loss, we saved its weights. Finally, we used these weights of the individual models for ensembling and performing classification on the test images. In particular, we applied the convolutional filters, padding, pooling filters, and strides at their default parameters from the original Resnet50 and Densenet201 networks (52; 53). The optimal values for each of the hyperparameters utilized in this analysis are shown in table 5.

**Table 5.** Hyperparameters in the Resnet50 and Densenet201 Model

| Hyperparameters | Resnet50 model | Densenet201 model |
| --- | --- | --- |
| Optimizer | Adam | Adam |
| Learning Rate | 0.001 | 0.001 |
| Loss Function | Binary Cross-entropy | Binary Cross-entropy |
| Batch Size | 32 | 32 |
| Epochs | 200 | 200 |
| Dropout | 0.2 | 0.2 |
| Input size | $256 \times 256 \times 3$ | $256 \times 256 \times 3$ |

### 5.3. Discussion of Resnet50 and Densenet201

Learning curves are used to represent the epoch-based assessment of a classifier’s performance over time. For both the training and validation sets, the learning curves are the accuracy and loss curves. Figure 11 illustrates a comparison of the Resnet50 and Densenet201 models’ accuracy and loss values. Each model has a varied accuracy and error rate, however the Resnet50 model outperforms Densenet201 model by obtaining an accuracy of 78.87% as shown in table 6.

**Table 6.** Evaluation metrics of Resnet50 and Densenet201 model without applying Empirical wavelet transform

| Metrics | Resnet50 | Densenet201 |
| --- | --- | --- |
| Accuracy | 78.87% | 77.46% |
| Specificity | 81.81% | 82.69% |
| Sensitivity | 68.75% | 63.15% |
| ROC AUC Score | 71.19% | 71.57% |

**Figure 11:**
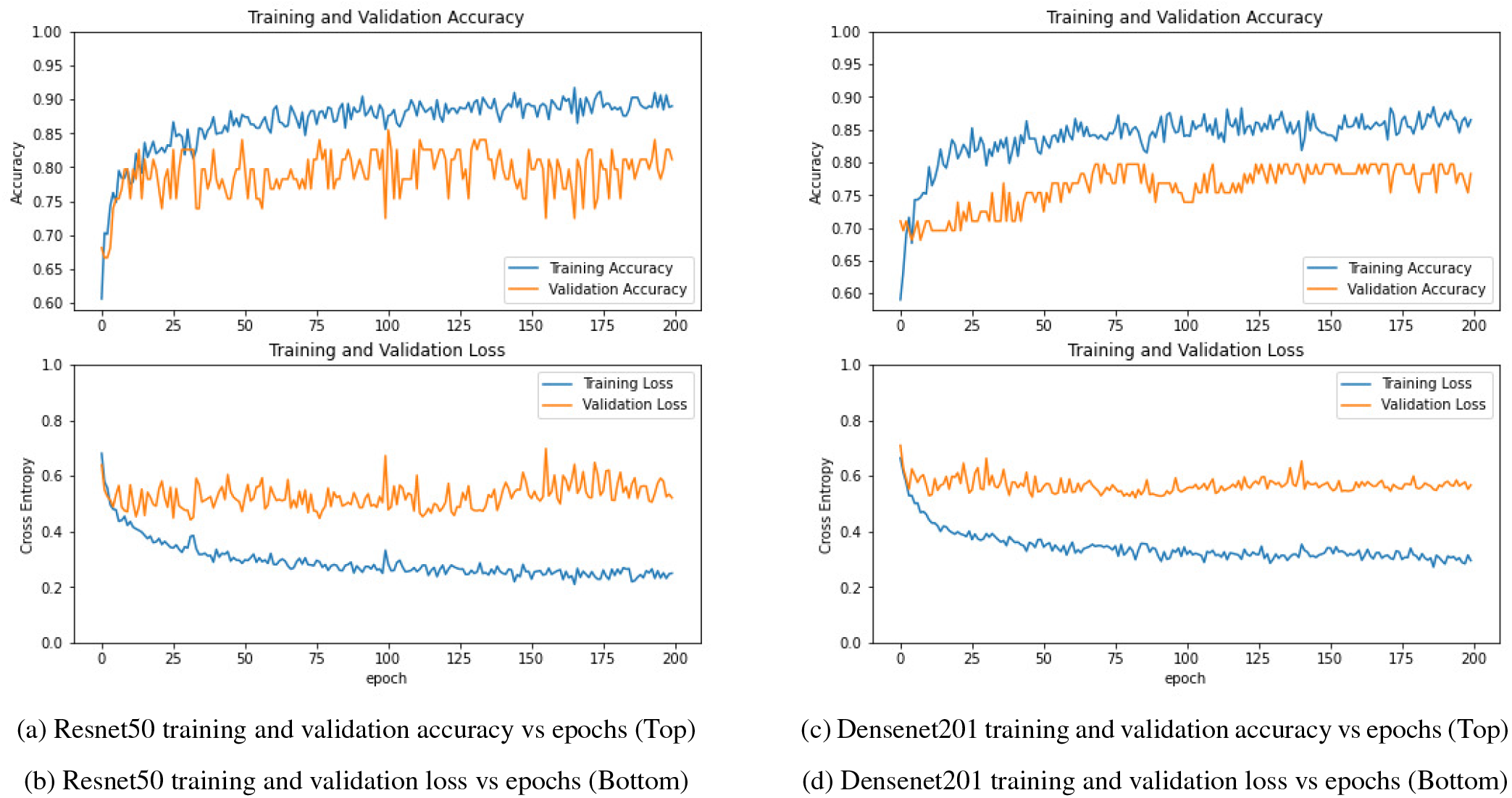
(a)Accuracy plot of Resnet50 model training over 200 epochs without EWT(b)Loss plot of Resnet50 model training over 200 epochs without EWT(c)Accuracy plot of Densenet201 model training over 200 epochs without EWT(d)Loss plot of Densenet201 model training over 200 epochs without EWT. It is evident that models trained without using EWT have a wider generalization gap between training and validation loss compared to models trained with EWT, which results in lower classification accuracy.

### 5.4. Discussion of Resnet50 and Densenet201 model results with EWT

Feature extraction is an importing aspect of deep learning. The images are sent to EWT module which decomposes the images into different sub-band images out of which the first sub-band image is fed to the Resnet50 and Densenet201 models for training. Significant improvements could be observed between figures 12 and 11, which depict the accuracy and loss curves. Thus, we say that EWT is highly effective as a feature extraction tool that helps in improving the performance of the models. Here also we observe that the Resnet50 model performs better than the Densenet201 model by obtaining an accuracy of 88.73%.

**Figure 12:**
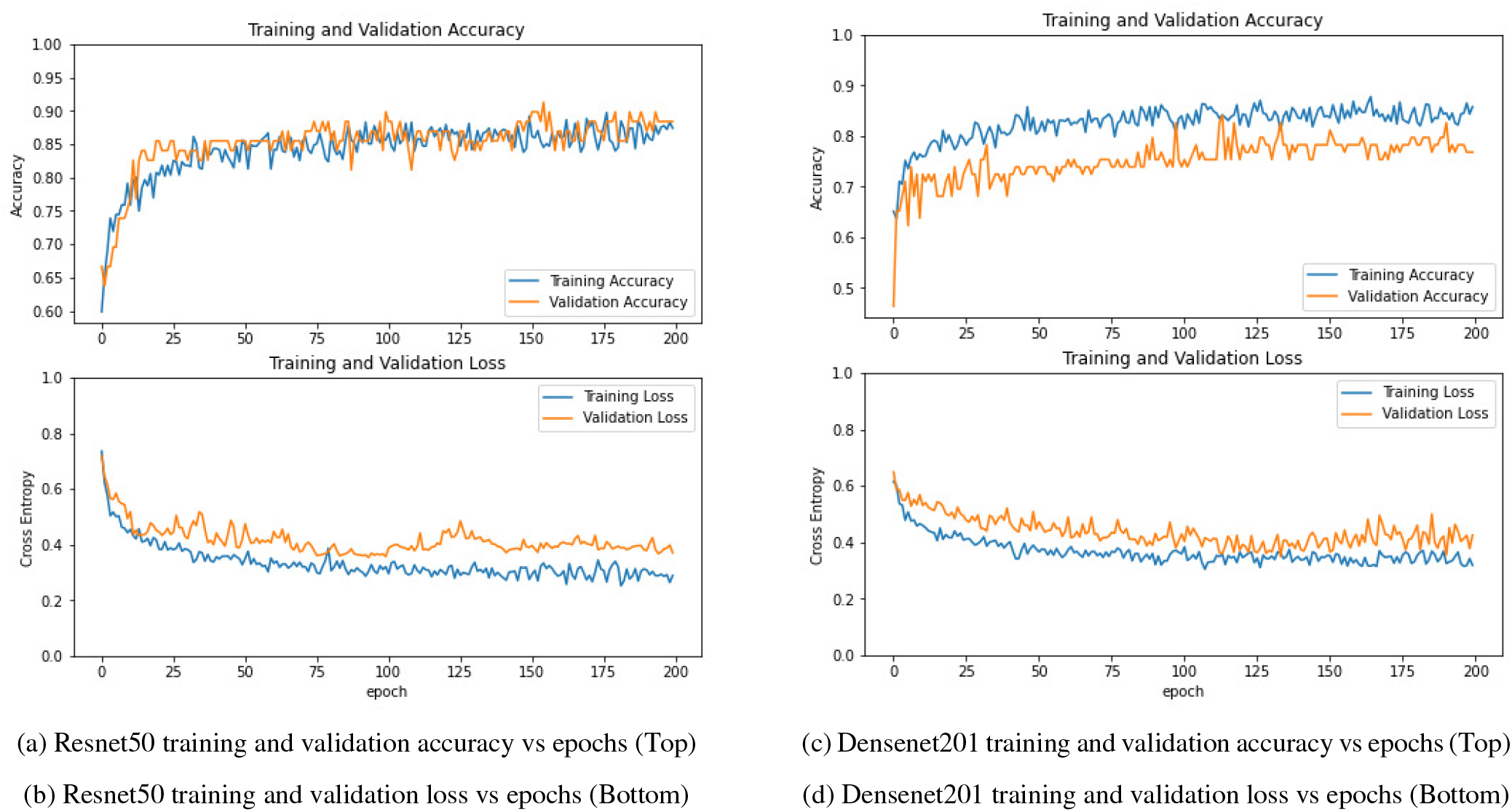
(a)Accuracy plot of Resnet50 model training over 200 epochs after EWT.The gap between training and validation accuracy is a clear indication that the model is not overfitting.(b)Loss plot of Resnet50 model training over 200 epochs after EWT. We can observe that the difference between the training and validation loss values narrows to a point of stability(c)Accuracy plot of Densenet201 model training over 200 epochs after EWT.The gap between the training and validation accuracy shows that the model slightly overfits compared to Resnet50 model(d)Loss plot of Densenet201 model training over 200 epochs after EWT. Similar to the Resnet50 model, the difference between the training and validation loss values stabilizes in this case as well.

### 5.5. Discussion of proposed ensemble model results

We proposed an efficient technique for ensembling our two customized architectures, Resnet50 and Densenet201 models by ensembling method. The weighted ensembling approach is carried out using both custom networks to increase the model’s stability and prediction accuracy. With the ensemble model, an accuracy of 92.00% was attained on the testing set. In table 6, 7, the model’s final observations are deduced. It showed that empirical wavelet transform acts as a powerful tool for feature extraction which helps in enhancing the performance of the model. Table 8 shows a few comparisons with previous works.

**Table 7.**
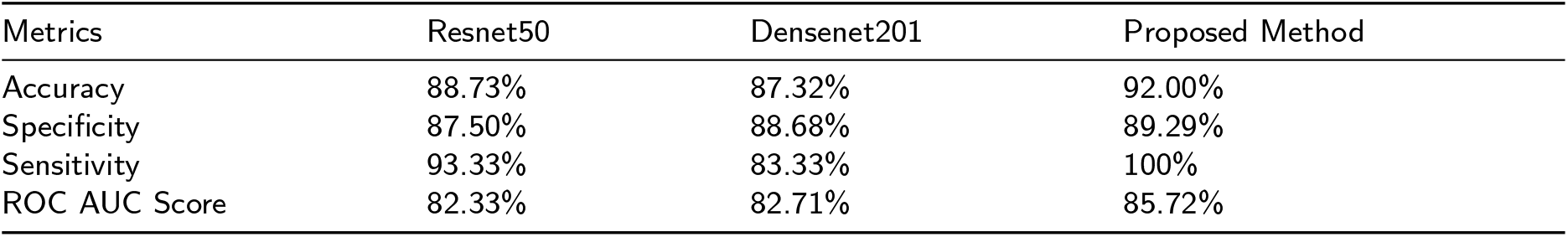
Evaluation metrics of the proposed model, Resnet50 and Densenet201 models by applying Empirical wavelet transform

| Metrics | Resnet50 | Densenet201 | Proposed Method |
| --- | --- | --- | --- |
| Accuracy | 88.73% | 87.32% | 92.00% |
| Specificity | 87.50% | 88.68% | 89.29% |
| Sensitivity | 93.33% | 83.33% | 100% |
| ROC AUC Score | 82.33% | 82.71% | 85.72% |

**Table 8.** Comparative analysis with previous research

| Research work | Classification method | Dataset source | Dataset ref. | Classification accuracy |
| --- | --- | --- | --- | --- |
| (74) | Resnet101 | Private | Not available | 78.30% |
| (75) | EfficientNet-B0 | Private | Not available | 85.00% |
| (37) | Deep learning | Private | Not available | 88.30% |
| (76) | Alexnet | Public | (70) | 90.06% |
| **Current study** | **Proposed method** | **Public** | **(70)** | **92.00%** |

## 6. Conclusion

In conclusion, we have a new empirical wavelet transform-based approach for feature extraction from oral histopathological images for binary image classification in this study. The EWT is a potential tool for performing medical image processing analysis due to its inherent adaptability. When the proposed method is contrasted with other approaches already in use, it becomes clear that the proposed methodology performs better. It is interesting that, out of all possible evaluated combinations, the first decomposed subband image obtained by the EWT decomposition technique yields the best results in terms of accuracy, sensitivity, and specificity. The same methodology can be used in future computer vision research to detect other diseases.

In this study, using our collected dataset, we presented an ensemble deep learning technique along with an empirical wavelet transform as a feature extraction tool for the categorization of oral cancer histopathological images. This study’s major goal was to efficiently classify images of cancer. We found that using the weighted average ensembling of two pre trained model performs better in comparison to individual models and thus a comparatively more robust model is achieved. When it comes to the classification of extremely complicated histopathological images of oral cancer, the proposed ensemble technique performs effectively.

The method’s main drawback is that it may be difficult to demonstrate the robustness and generalizability of the approach if the data are not readily available (77). The dataset employed in this study has just 696 images, whereas the more efficient deep learning models require larger datasets to be trained on for maximum performance depending on the complexity of the problem. Deep learning models fundamentally work best with a very large database. Since ImageNet contains 14 million images, we were required to use transfer learning models that were fine-tuned using the histopathological images during our study after being pretrained on ImageNet. The expansion of our dataset and the incorporation of images for multi-class classification issues are two future directions for this study. Future work also needs to incorporate other pre-trained deep-learning models. It will be remarkable to see how the proposed ensemble criteria perform when applied to histopathological images of other tumors, including breast, cervical, and lung cancer.

## Data Availability

All data produced are available online at
Rahman, T. Y., Mahanta, L. B., Das, A. K.&Sarma, J. D. Histopathological imaging database for oral cancer analysis. Data in brief 29,
105114 (2020).

https://www.ncbi.nlm.nih.gov/pmc/articles/PMC6994517/

## CRediT authorship contribution statement

### Bhaswati Singha Deo

Developed the model from the concept, Developed the code, Generated the results, Wrote the manuscript. **Mayukha Pal:** Conceived the idea and conceptualized it, Developed the methodology from the concept, Reviewed the manuscript and mentored the work. **Prasanta K. Panigrahi:** Reviewed the manuscript and mentored the work. **Asima Pradhan:** Developed the analysis methodology, Reviewed the manuscript and mentored the work.

## Acknowledgement

This research received no specific support from governmental, private, or non-profit funding agencies. The content and writing of the paper are solely the responsibility of the authors. Bhaswati Singha Deo is thankful to IIT Kanpur for the Institute fellowship. The author MPal wishes to thank ABB Ability Innovation Centre, India for their support in this work.

## References

[1] Li, L., Yin, Y., Nan, F. & Ma, Z. Circ_lpar3 promotes the progression of oral squamous cell carcinoma (oscc). Biochemical and Biophysical Research Communications 589, 215–222 (2022).

[2] Saba, T. Recent advancement in cancer detection using machine learning: Systematic survey of decades, comparisons and challenges. Journal of Infection and Public Health 13 (9), 1274–1289 (2020).

[3] World cancer research fund international, https://www.wcrf.org/cancer-trends/mouth-and-oral-cancer-statistics/.

[4] Chakraborty, D., Natarajan, C. & Mukherjee, A. Advances in oral cancer detection. Advances in clinical chemistry 91, 181–200 (2019).

[5] Eckert, A. W., Kappler, M., Große, I., Wickenhauser, C. & Seliger, B. Current understanding of the hif-1-dependent metabolism in oral squamous cell carcinoma. International journal of molecular sciences 21 (17), 6083 (2020).

[6] Ghosh, A. et al. Deep reinforced neural network model for cyto-spectroscopic analysis of epigenetic markers for automated oral cancer risk prediction. Chemometrics and Intelligent Laboratory Systems 224, 104548 (2022).

[7] Deif, M. A. & Hammam, R. E. Skin lesions classification based on deep learning approach. Journal of Clinical Engineering 45 (3), 155–161 (2020).

[8] Kong, J. et al. Computer-aided evaluation of neuroblastoma on whole-slide histology images: Classifying grade of neuroblastic differentiation. Pattern Recognition 42 (6), 1080–1092 (2009).

[9] Santana, M. F. & Ferreira, L. C. L. Diagnostic errors in surgical pathology. Jornal Brasileiro de Patologia e Medicina Laboratorial 53, 124–129 (2017).

[10] Kaladhar, D., Chandana, B. & Kumar, P. B. Predicting cancer survivability using classification algorithms. International Journal of Research and Reviews in Computer Science 2 (2), 340 (2011).

[11] Mandal, A., Tiwari, Y., Panigrahi, P. K. & Pal, M. Physics aware analytics for accurate state prediction of dynamical systems. Chaos, Solitons & Fractals 164, 112670 (2022).

[12] Kann, B. H. et al. Pretreatment identification of head and neck cancer nodal metastasis and extranodal extension using deep learning neural networks. Scientific reports 8 (1), 1–11 (2018).

[13] Petit, O., Thome, N. & Soler, L. Iterative confidence relabeling with deep convnets for organ segmentation with partial labels. Computerized Medical Imaging and Graphics 91, 101938 (2021).

[14] Deif, M. A., Solyman, A. A., Alsharif, M. H. & Uthansakul, P. Automated triage system for intensive care admissions during the covid-19 pandemic using hybrid xgboost-ahp approach. Sensors 21 (19), 6379 (2021).

[15] Echle, A. et al. Deep learning in cancer pathology: a new generation of clinical biomarkers. British journal of cancer 124 (4), 686–696 (2021).

[16] Karthik, R., Menaka, R. & Siddharth, M. Classification of breast cancer from histopathology images using an ensemble of deep multiscale networks. Biocybernetics and Biomedical Engineering 42 (3), 963–976 (2022).

[17] Kareem, S. A., Pozos-Parra, P. & Wilson, N. An application of belief merging for the diagnosis of oral cancer. Applied Soft Computing 61, 1105–1112 (2017).

[18] Duggento, A., Conti, A., Mauriello, A., Guerrisi, M. & Toschi, N. Deep computational pathology in breast cancer, Vol. 72, 226–237 (Elsevier, 2021).

[19] Goldenberg, S. L., Nir, G. & Salcudean, S. E. A new era: artificial intelligence and machine learning in prostate cancer. Nature Reviews Urology 16 (7), 391–403 (2019).

[20] Wang, S. et al. Artificial intelligence in lung cancer pathology image analysis. Cancers 11 (11), 1673 (2019).

[21] Majumdar, M. & Gayen, T. in Computer aided segmentation of oral mucosa to detect cancer 179–190 (CRC Press, 2022).

[22] Krishnan, M., Acharya, U., Chakraborty, C. & Ray, A. Automated diagnosis of oral cancer using higher order spectra features and local binary pattern: A comparative study. Technology in cancer research & treatment 10 (5), 443–455 (2011).

[23] Prabhakar, S. K. & Rajaguru, H. Performance analysis of linear layer neural networks for oral cancer classification, 1–4 (IEEE, 2017).

[24] Patra, R., Chakraborty, C. & Chatterjee, J. Textural analysis of spinous layer for grading oral submucous fibrosis. Int J Comput Appl 47, 975–8887 (2012).

[25] Krishnan, M. M. R. et al. Textural characterization of histopathology-ical images for oral sub-mucous fibrosis detection. Tissue and Cell 43 (5), 318–330 (2011).

[26] Thomas, B., Kumar, V. & Saini, S. Texture analysis based segmentation and classification of oral cancer lesions in color images using ann, 1–5 (IEEE, 2013).

[27] Rahman, T., Mahanta, L., Chakraborty, C., Das, A. & Sarma, J. Textural pattern classification for oral squamous cell carcinoma. Journal of microscopy 269 (1), 85–93 (2018).

[28] Rahman, T. Y., Mahanta, L. B., Das, A. K. & Sarma, J. D. Automated oral squamous cell carcinoma identification using shape, texture and color features of whole image strips. Tissue and Cell 63, 101322 (2020).

[29] Alabi, R. O. et al. Machine learning in oral squamous cell carcinoma: Current status, clinical concerns and prospects for future—a systematic review. Artificial Intelligence in Medicine 115, 102060 (2021)

[30] Sun, M.-L. et al. Application of machine learning to stomatology: a comprehensive review. IEEE Access 8, 184360–184374 (2020).

[31] Alkhadar, H., Macluskey, M., White, S., Ellis, I. & Gardner, A. Comparison of machine learning algorithms for the prediction of five-year survival in oral squamous cell carcinoma. Journal of Oral Pathology & Medicine 50 (4), 378–384 (2021).

[32] Mehmood, S. et al. Malignancy detection in lung and colon histopathology images using transfer learning with class selective image processing. IEEE Access 10, 25657–25668 (2022).

[33] Krizhevsky, A., Sutskever, I. & Hinton, G. E. Imagenet classification with deep convolutional neural networks. Communications of the ACM 60 (6), 84–90 (2017).

[34] Srinidhi, C. L., Ciga, O. & Martel, A. L. Deep neural network models for computational histopathology: A survey. Medical Image Analysis 67, 101813 (2021).

[35] Das, N., Hussain, E. & Mahanta, L. B. Automated classification of cells into multiple classes in epithelial tissue of oral squamous cell carcinoma using transfer learning and convolutional neural network. Neural Networks 128, 47–60 (2020).

[36] Folmsbee, J., Liu, X., Brandwein-Weber, M. & Doyle, S. Active deep learning: Improved training efficiency of convolutional neural networks for tissue classification in oral cavity cancer, 770–773 (IEEE, 2018).

[37] Aubreville, M. et al. Automatic classification of cancerous tissue in laserendomicroscopy images of the oral cavity using deep learning. Scientific reports 7 (1), 1–10 (2017).

[38] Kassani, S. H., Kassani, P. H., Wesolowski, M. J., Schneider, K. A. & Deters, R. Classification of histopathological biopsy images using ensemble of deep learning networks. arXiv preprint arXiv:1909.11870 (2019).

[39] Simonyan, K. & Zisserman, A. Very deep convolutional networks for large-scale image recognition. arXiv preprint arXiv:1409.1556 (2014).

[40] Szegedy, C. et al. Going deeper with convolutions, 1–9 (2015).

[41] Gaur, D., Folz, J. & Dengel, A. Training deep neural networks without batch normalization. arXiv preprint arXiv:2008.07970 (2020).

[42] Gilles, J. Empirical wavelet transform. IEEE transactions on signal processing 61 (16), 3999–4010 (2013).

[43] Gilles, J., Tran, G. & Osher, S. 2d empirical transforms. wavelets, ridgelets, and curvelets revisited. SIAM Journal on Imaging Sciences 7 (1), 157–186 (2014).

[44] Aneesh, C., Kumar, S., Hisham, P. & Soman, K. Performance comparison of variational mode decomposition over empirical wavelet transform for the classification of power quality disturbances using support vector machine. Procedia Computer Science 46, 372–380 (2015).

[45] Thirumala, K., Umarikar, A. C. & Jain, T. Estimation of single-phase and three-phase power-quality indices using empirical wavelet transform. IEEE Transactions on power delivery 30 (1), 445–454 (2014).

[46] Li, Y., Xue, B., Hong, H. & Zhu, X. Instantaneous pitch estimation based on empirical wavelet transform, 250–253 (IEEE, 2014).

[47] Hu, J., Wang, J. & Ma, K. A hybrid technique for short-term wind speed prediction. Energy 81, 563–574 (2015).

[48] Kedadouche, M., Thomas, M. & Tahan, A. A comparative study between empirical wavelet transforms and empirical mode decomposition methods: Application to bearing defect diagnosis. Mechanical Systems and Signal Processing 81, 88–107 (2016).

[49] Sharma, R. R., Kalyani, A. & Pachori, R. B. An empirical wavelet transform-based approach for cross-terms-free wigner–ville distribution. Signal, Image and Video Processing 14 (2), 249–256 (2020).

[50] Sharma, R. R., Kumar, M. & Pachori, R. B. Joint time-frequency domain-based cad disease sensing system using ecg signals. IEEE Sensors Journal 19 (10), 3912–3920 (2019).

[51] Huang, N. E. et al. The empirical mode decomposition and the hilbert spectrum for nonlinear and non-stationary time series analysis. Proceedings of the Royal Society of London. Series A: mathematical, physical and engineering sciences 454 (1971), 903–995 (1998).

[52] He, K., Zhang, X., Ren, S. & Sun, J. Deep residual learning for image recognition, 770–778 (2016).

[53] Huang, G., Liu, Z., Van Der Maaten, L. & Weinberger, K. Q. Densely connected convolutional networks, 4700–4708 (2017).

[54] Deng, J. et al. Imagenet: A large-scale hierarchical image database, 48–255 (Ieee, 2009).

[55] Shin, H.-C. et al. Deep convolutional neural networks for computeraided detection: Cnn architectures, dataset characteristics and transfer learning. IEEE transactions on medical imaging 35 (5), 1285–1298 (2016).

[56] Byra, M. et al. Knee menisci segmentation and relaxometry of 3d ultrashort echo time cones mr imaging using attention u-net with transfer learning. Magnetic resonance in medicine 83 (3), 1109–1122 (2020).

[57] Tang, X., Du, B., Huang, J., Wang, Z. & Zhang, L. On combining active and transfer learning for medical data classification. IET Computer Vision 13 (2), 194–205 (2019).

[58] Zeng, M. et al. Automatic icd-9 coding via deep transfer learning. Neurocomputing 324, 43–50 (2019).

[59] Dietterich, T. G. Ensemble methods in machine learning, 1–15 (Springer, 2000).

[60] Brown, G., Wyatt, J., Harris, R. & Yao, X. Diversity creation methods: a survey and categorisation. Information fusion 6 (1), 5–20 (2005).

[61] Pal, M., Tiwari, Y., Reddy, T. V., Parisineni, S. R. A. & Panigrahi, P. K. An integrative method for covid-19 patients classification from chest x-ray using deep learning network with image visibility graph as feature extractor. medRxiv (2021).

[62] Khaki, S. & Wang, L. Crop yield prediction using deep neural networks. Frontiers in plant science 10, 621 (2019).

[63] Taieb, S. B. & Hyndman, R. J. A gradient boosting approach to the kaggle load forecasting competition. International journal of forecasting 30 (2), 382–394 (2014).

[64] Yu, H.-F. et al. Feature engineering and classifier ensemble for kdd cup 2010 (2010).

[65] Breiman, L. Bagging predictors. Machine learning 24 (2), 123–140 (1996).

[66] Freund, Y. Boosting a weak learning algorithm by majority. Information and computation 121 (2), 256–285 (1995).

[67] Wolpert, D. H. Stacked generalization. Neural networks 5 (2), 241–259 (1992).

[68] Drucker, H., Cortes, C., Jackel, L. D., LeCun, Y. & Vapnik, V. Boosting and other ensemble methods. Neural Computation 6 (6), 1289–1301 (1994).

[69] Large, J., Lines, J. & Bagnall, A. A probabilistic classifier ensemble weighting scheme based on cross-validated accuracy estimates. Data mining and knowledge discovery 33 (6), 1674–1709 (2019).

[70] Rahman, T. Y., Mahanta, L. B., Das, A. K. & Sarma, J. D. Histopathological imaging database for oral cancer analysis. Data in brief 29, 105114 (2020).

[71] Goodfellow, I., Bengio, Y. & Courville, A. Deep learning (MIT press, 2016).

[72] Kingma, D. P. & Ba, J. Adam: A method for stochastic optimization. arXiv preprint arXiv:1412.6980 (2014).

[73] Srivastava, N., Hinton, G., Krizhevsky, A., Sutskever, I. & Salakhutdinov, R. Dropout: a simple way to prevent neural networks from overfitting. The journal of machine learning research 15 (1), 1929–1958 (2014).

[74] Welikala, R. A. et al. Automated detection and classification of oral lesions using deep learning for early detection of oral cancer. IEEE Access 8, 132677–132693 (2020).

[75] Jubair, F. et al. A novel lightweight deep convolutional neural network for early detection of oral cancer. Oral Diseases 28 (4), 1123–1130 (2022).

[76] Rahman, A.-u. et al. Histopathologic oral cancer prediction using oral squamous cell carcinoma biopsy empowered with transfer learning. Sensors 22 (10), 3833 (2022).

[77] Sengupta, N., Sarode, S. C., Sarode, G. S. & Ghone, U. Scarcity of publicly available oral cancer image datasets for machine learning research. Oral Oncology 126, 105737 (2022).

